# Drug-drug interaction identification using large language models

**DOI:** 10.64898/2025.12.03.25341549

**Authors:** Kaitlin Blotske, Xingmeng Zhao, Kelli Henry, Yanjun Gao, Adeleine Tilley, Moriah Cargile, Brian Murray, Susan E. Smith, Erin F. Barreto, Seth Bauer, Sunghwan Sohn, Tianming Liu, Tell Bennett, Mitch Cohen, Hongjian Zhou, Fenglin Liu, David A. Clifton, Andrea Sikora

## Abstract

**Background:** The purpose of this study was to develop a series of benchmarks specifically for large language model (LLM) performance in the identification of drug-drug interactions (DDIs).

**Methods:** We evaluated six LLMs using a clinician-designed dataset of 750 DDI scenarios. Tasks included: (1) a pointwise two-drug classification task (2) a pairwise three-drug discrimination task and (3) a listwise 4–6 drug selection task. Zero-shot prompting was applied and performance was assessed using precision, recall, F1 score, accuracy, and self-consistency.

**Results:** Model performance varied by task structure and interaction severity. GPT-5-Chat demonstrated the highest recall and F1 score in the pointwise task (80.7% and 80.2%), whereas DrugGPT achieved superior accuracy in the pairwise (93.7%) and listwise task (92.6%). All models had improved performance with more severe compared to less severe DDIs (accuracy 72.0-99.2% vs. 58.3-86.7%). All models tended to have higher self-consistency in the pairwise and listwise experiments compared to the pointwise experiment (68.8-93.6% vs. 31.0-74.2%). No model exhibited uniformly high reliability across all tasks.

**Conclusions:** Current LLMs show inconsistent capabilities in identifying DDIs. Performance degrades as reasoning space expands with limited stability. These findings emphasize the need for multi-format evaluation frameworks and reliability-aware assessment when considering LLMs for medication-safety applications.

## Background

Clinical natural language processing (cNLP), particularly in the form of large language models (LLMs), has completed impressive feats in the healthcare space, and great interest exists for how these new technologies can improve healthcare, including medication safety. Importantly though, a recent Federal Food and Drug Administration (FDA) statement was published regarding artificial intelligence (AI) regulation stating: “The sheer volume of these changes and their impact also suggests the need for industry and other external stakeholders to ramp up assessment and quality management of AI across the larger ecosystem beyond the remit of the FDA…all involved sectors will need to attend to AI with the care and rigor this potentially transformative technology merits.”^1^ As such, careful probing and documentation of LLM performance is imperative for safe and efficacious deployment for medication-related tasks.

Previous studies that have evaluated LLMs for drug-drug interaction (DDI) tasks demonstrated initial promise but also identified errors that have potential to worsen patient safety without further methodological development.^2–17^ This is particularly true because LLMs would be replacing a high-performing standard of care (i.e., rules-based computer software that can reliably identify any DDI in its bank).^2–17^ To date, no studies have focused on benchmarking performance with the goal that LLMs could be said to have safe and effective performance for clinical use. Notably, the generated datasets that have been used prior have been reviewed and demonstrated inaccuracies or lack of clinical relevance.^18,19^ For example, DDI-Corpus, one of the largest DDI databases with over 5,000 DDIs, was developed using DrugBank and Medline abstracts but had no clinical validation resulting in a large portion of the listed DDIs containing conflicting information (for example, nonsteroidal anti-inflammatory drugs [NSAIDs] and aspirin listed as “no relation” and “advise” depending on the referenced article) or incorrect information (for example, “warfarin” and “coumadin” listed as having no relation).^18^ Additionally, many of the medications included are outdated and not available on the market.^18^ Several LLMs have used DDI-Corpus for training and evaluation of DDIs, but caution is warranted for the application of their results given the lack of clinical validity in the training dataset.^20^

The purpose of this study was to develop a series of benchmarks specifically for LLM performance in the identification of DDIs using a curated clinician-designed dataset of clinically-relevant DDIs.

## Methods

### Study design

We designed a three-part evaluation framework to assess how LLMs identify DDIs across tasks that mirror real-world clinical decision-making. Six models were evaluated: GPT-5-Chat, GPT-4o-mini, MedGemma-27B, LLaMA3-70B, Qwen3-32B, and DrugGPT. To capture different aspects of DDI reasoning, we implemented three complementary experiments with nine runs each: pointwise, pairwise, and listwise DDI identification.

*1. Pointwise* refers to a two-drug experiment consisting of interacting (Category C, D, and X) and non-interacting (Category A) drug pairs where the LLM answered interaction present or not present.
*2. Pairwise* refers to a three-drug experiment that included one target drug and two candidate drugs, exactly one of which produced a known interaction and asked the LLM to provide the interacting pair.
*3. Listwise* refers to 4–6 drug experiment that contained a short medication list with a single interacting drug pair within each list and asked the LLM to identify the interacting pair.

In this framework, *pointwise* evaluation assesses the correctness of a single candidate in isolation (e.g., verifying whether a proposed DDI category is clinically valid), *pairwise* evaluation requires the model to discriminate between two competing alternatives (e.g., selecting the more accurate interaction label or explanation), and *listwise* evaluation extends this comparison to multiple candidates simultaneously, requiring ranking or selection among several possible options, thereby capturing different dimensions of decision consistency and comparative reasoning. These formats have been adopted in recent LLM-as-judge literature and support assessment of single-item correctness, comparative discrimination among alternatives, and multi-option selection, providing a more complete view of model behavior.^21^ Further, this structure follows the logic of “multi-format judgment” introduced in tool-integrated LLM judge frameworks and has been shown to reveal different failure patterns that would be missed using only a single evaluation mode.^22^

This project was reviewed and approved by the University of Colorado Institutional Review Board (COMIRB #25-1631). All methods were performed in accordance with the ethical standards of the Helsinki Declaration of 1975.^23^ This evaluation followed the transparent reporting of a multivariable model for individual prognosis or diagnosis (TRIPOD–LLM) extension reporting frameworks, as applicable (**Supplemental Appendix**).^24^

### Dataset Development

A clinician-curated dataset of 250 unique DDI scenarios for the three experiments was developed. Interaction labels were sourced from LexiDrug, which provides standardized definitions for Categories A (no known interaction), B (no action needed), C (monitor therapy), D (consider therapy modification), and X (avoid combination).^25^ Dataset construction followed the clinical-validation process used in prior pharmacology-oriented LLM research: all cases were independently reviewed by three board-certified clinical pharmacists, and only scenarios with full agreement were included for this analysis.^3,6,10,16,17,20,26,27^ This ensured that the benchmark reflected *clinically relevant* DDIs with accurate severity classifications. These datasets are posted on Github: https://github.com/sikora07/AIChemist. Characteristics of each dataset are summarized in **Table 1**.

**Table 1.** Description of datasets.

|  | **Drug Pair Experiment**<br>**(n=250)** | **3 Drug Combination**<br>**Experiment (n=250)** | **4-6 Drug Combination**<br>**Experiment (n=250)** |
| --- | --- | --- | --- |
| **1 Drug Interaction**<br>**Present** | 125 (50) | 250 (100) | 250 (100) |
| **Category A** | 125 (50) | 0 (0) | 0 (0) |
| **Category B** | 0 (0) | 0 (0) | 0 (0) |
| **Category C** | 40 (16) | 82 (32.8) | 81 (32.4) |
| **Category D** | 45 (18) | 78 (31.2) | 77 (30.8) |
| **Category X** | 40 (16) | 90 (36) | 92 (36.8) |
| **Number of Medications**<br>**Per Prompt** | 2 (2-2) | 3 (3-3) | 4 (4-5) |

Each experiment contained a total of 250 groups with unique medication scenarios. Experiment “drug-pair” is the only assessment containing 50% of groups with “negative” or “no known interactions”. The remainder of experiments contained drug-drug interaction pairs within C, D, or X interaction categories.
All results as n (%) or median (IQR)
Drug interaction categories are defined as follows:
Category X — Avoid combination: This drug pair should generally not be used together because the clinical risk outweighs any potential benefit.
Category D — Consider therapy modification: The interaction is clinically relevant, and modification of therapy (dose adjustment, substitution, or precautions) should be considered.
Category C — Monitor therapy: An interaction is present. The combination may be used but requires monitoring by a healthcare professional.
Category B — No action needed: An interaction may exist, but no clinical intervention is required.
Category A — No known interaction: There is no known documented interaction between the two drugs.

### Prompting Procedure

All models were evaluated using standardized zero-shot prompts tailored to the specific structure of each task (**Table 2**). To minimize parsing errors and hallucinations, prompts incorporated rigorous clinical definitions derived from standard interaction categories (e.g., Category A-X) and enforced strict output formats, specifically, a binary ‘A’/’B’ selection or a structured JavaScript Object Notation (JSON) format.^25^

**Table 2.** Input and output prompts for three drug-drug interaction benchmark experiments.

| Benchmark | Input | Output |
| --- | --- | --- |
| **DDI Pair** | You are a clinical pharmacist specializing in drug-drug interactions. Drug-drug interactions may occur when one medication alters the clinical effect of another (pharmacodynamic interaction) or affects its absorption, distribution, metabolism, or excretion (pharmacokinetic interaction).<br>Drug interaction categories are defined as follows:<br>Category X — Avoid combination: This drug pair should generally not be used together because the clinical risk outweighs any potential benefit.<br>Category D — Consider therapy modification: The interaction is clinically relevant, and modification of therapy (dose adjustment, substitution, or precautions) should be considered.<br>Category C — Monitor therapy: An interaction is present. The combination may be used but requires monitoring by a healthcare professional.<br>Category B — No action needed: An interaction may exist, but no clinical intervention is required.<br>Category A — No known interaction: There is no known documented interaction between the two drugs.<br>Your job is to evaluate whether the drug pair has been appropriately categorized into the correct drug-drug interaction category according to these definitions.<br>Drug Pair: {drug_pair}<br>Proposed Classification: {proposed_category}: {proposed_action}<br>Question: Is this classification correct?<br>A: "CORRECT"<br>B: "INCORRECT"<br>Just return "A" or "B", with no text around it. | A or B |
| **DDI 3 Drug Combination** | You are evaluating drug-drug interactions.<br>For the given drug, which drug (A or B) results in a category C, D, or X interaction?<br>Drug interaction categories are defined as follows:<br>Category X — Avoid combination: This drug pair should generally not be used together because the clinical risk outweighs any potential benefit.<br>Category D — Consider therapy modification: The interaction is clinically relevant, and modification of therapy (dose adjustment, substitution, or precautions) should be considered.<br>Category C — Monitor therapy: An interaction is present. The combination may be used but requires monitoring by a healthcare professional. | A or B |
|  | Drug Interaction Category Definition in Use: Category {category_label}<br>Question: Which drug causes a Category {category_label} interaction with {target_drug}?<br>A: candidate_drug_a<br>B: candidate_drug_b<br>Just return "A" or "B" with no text around it. |  |
| **DDI 4-6 Drug Combination** | You are a clinical pharmacist specializing in drug-drug interactions.<br>You will be given a list of medications taken together. Identify two medications that have a category C, D, or X drug interaction. Only identify one drug pair in JSON format. Do not estimate confidence or include any explanation.<br>Drug interaction categories are defined as follows:<br>Category X — Avoid combination: This drug pair should generally not be used together because the clinical risk outweighs any potential benefit.<br>Category D — Consider therapy modification: The interaction is clinically relevant, and modification of therapy (dose adjustment, substitution, or precautions) should be considered.<br>Category C — Monitor therapy: An interaction is present. The combination may be used but requires monitoring by a healthcare professional.<br>Medication List: {medication_lis<br>Format:<br>`{ "interactions": [ ["<drug_x>", "<drug_y>"] ] }`<br>Do not include any text before or after the JSON. Do not wrap the JSON in markdown code blocks. Return only the raw JSON object. | Drug X<br>Drug Y |

DDI: drug-drug interaction

To ensure the reliability of our results and account for the stochastic nature of LLMs, we employed a robust repeated-measures design. Each unique prompt was queried across nine independent runs under identical sampling parameters (temperature=0.7). This repetition allowed us to distinguish between stable clinical reasoning and stochastic variation, filtering out "lucky guesses" from robust knowledge. To further mitigate artifacts such as position bias (the tendency of LLMs to prefer the first option presented), we implemented a label shuffling mechanism. Across the nine runs, the assignment of semantic labels (e.g., Correct/Incorrect for pointwise tasks or Candidate A/Candidate B for discrimination tasks) to the output tokens (‘A’ or ‘B’) was randomized. This process ensured that reported accuracy reflects true semantic understanding rather than token preference. For the pointwise evaluation task, we adopted a balanced negative sampling strategy in which a negative example represents a counterfactual clinical claim, meaning a valid drug pair is paired with an incorrect drug–drug interaction (DDI) category (for example, assigning a Category X label to a pair whose verified category is Category C). Clinically, such counterfactuals reflect misleading interaction assessments that may cause inappropriate avoidance of safe combinations or failures to identify high-risk contraindications. For each ground-truth positive instance, we procedurally generated a matched negative instance by randomly selecting an incorrect category for the same pair, resulting in a one-to-one positive to negative distribution. This design requires the model not only to recognize correct interaction labels but also to detect and reject clinically implausible or unsafe categorizations, which tests its ability to identify false clinical assertions, recognize contradictions with established pharmacological evidence, and minimize the effects of acquiescence bias.

### Performance Metrics

Performance metrics were selected to align with the objectives of each experiment. All metrics were computed at the clinical case level, where a *case* refers to the full input provided to the model (e.g., a single drug pair, a three-drug prompt, or a 4–6 drug list). Each case yielded exactly one prediction and therefore contributed a single accuracy or error outcome, regardless of how many model tokens or intermediate steps were generated.

For the two-drug classification task (pointwise setting), we report accuracy, precision, recall, and F1. For the three-drug (pairwise) and multi-drug (listwise) tasks, accuracy was defined as selecting the correct interacting drug or the correct interacting drug pair for each complete case. Across all experiments, we also measured self-consistency to assess how reliably each model produced correct answers across repeated runs. Self-consistency was defined as number of runs that produced the correct answer divided by the total number of runs for that case. This metric captures whether a model consistently arrives at the correct answer when asked multiple times. A model that only occasionally produces the correct response will have low self-consistency even if its single-run accuracy appears acceptable, making this measure important for evaluating reliability in medication-safety tasks.

### Statistical Analysis

We evaluated model performance using each full clinical case as the unit of analysis. To quantify uncertainty in the reported metrics, we computed 95% confidence intervals using the percentile bootstrap with 1,000 resamples. In each bootstrap iteration, we drew a new sample of complete clinical cases with replacement and recalculated the metric of interest. We did not resample individual model outputs from the same case. This approach ensures that the confidence intervals reflect how model performance might change when applied to new DDI cases, rather than capturing randomness from repeated generations of the same prompt. Given the hypothesis generating nature of this exploration, no attempt was made to calculate sample size. All analyses were performed using Hugging Face package.^28^

## Results

We evaluated six large language models, GPT-5-Chat, GPT-4o-mini, MedGemma-27B, LLaMA3-70B, Qwen3-32B, and DrugGPT across three DDI tasks. Performance for each task is summarized in **Table 3**. Overall, model behavior differed substantially across pointwise, pairwise, and listwise settings, with no model demonstrating uniformly superior performance. Patterns across experiments highlight both strengths and persistent limitations in clinical reasoning and stability.

**Table 3.**
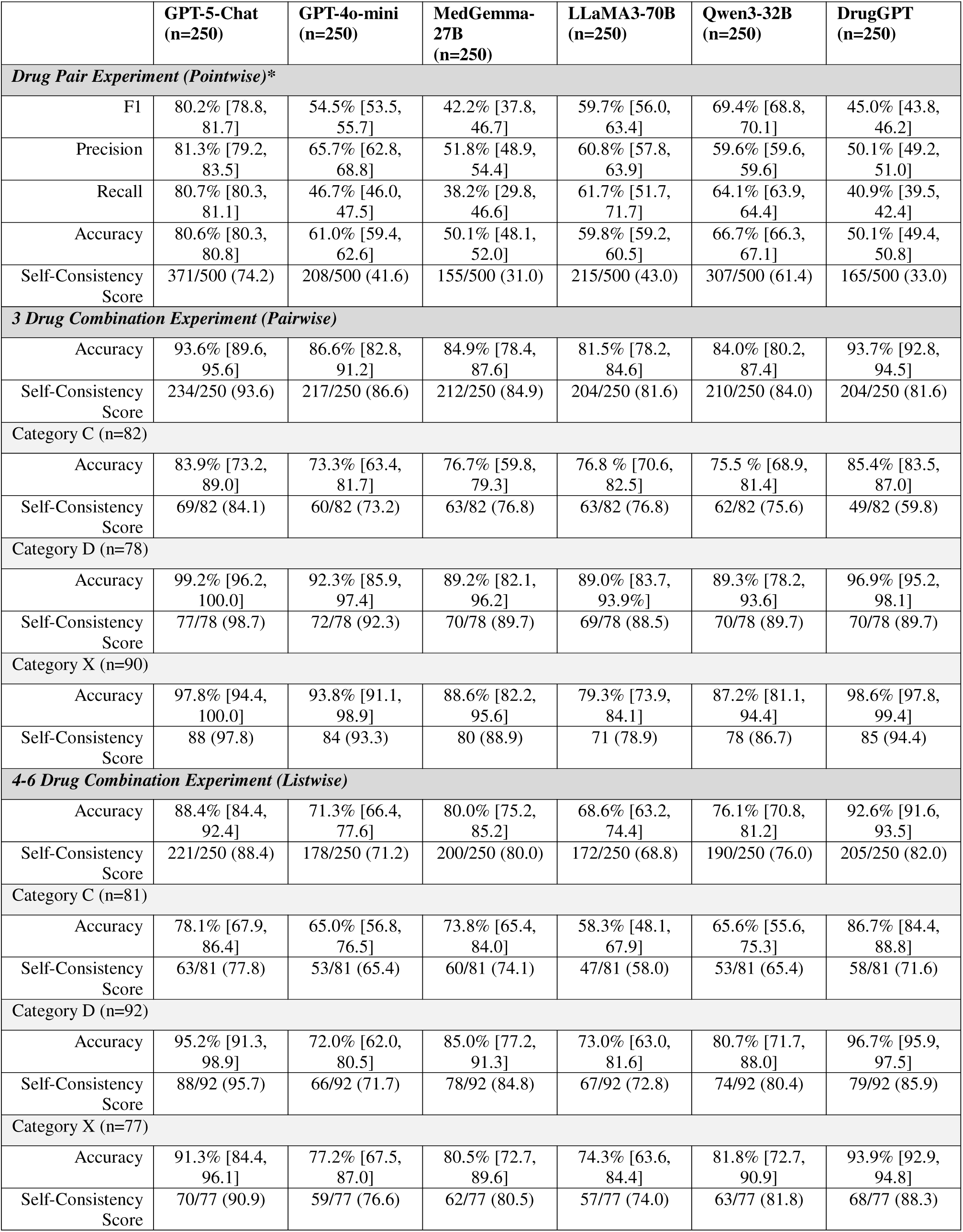
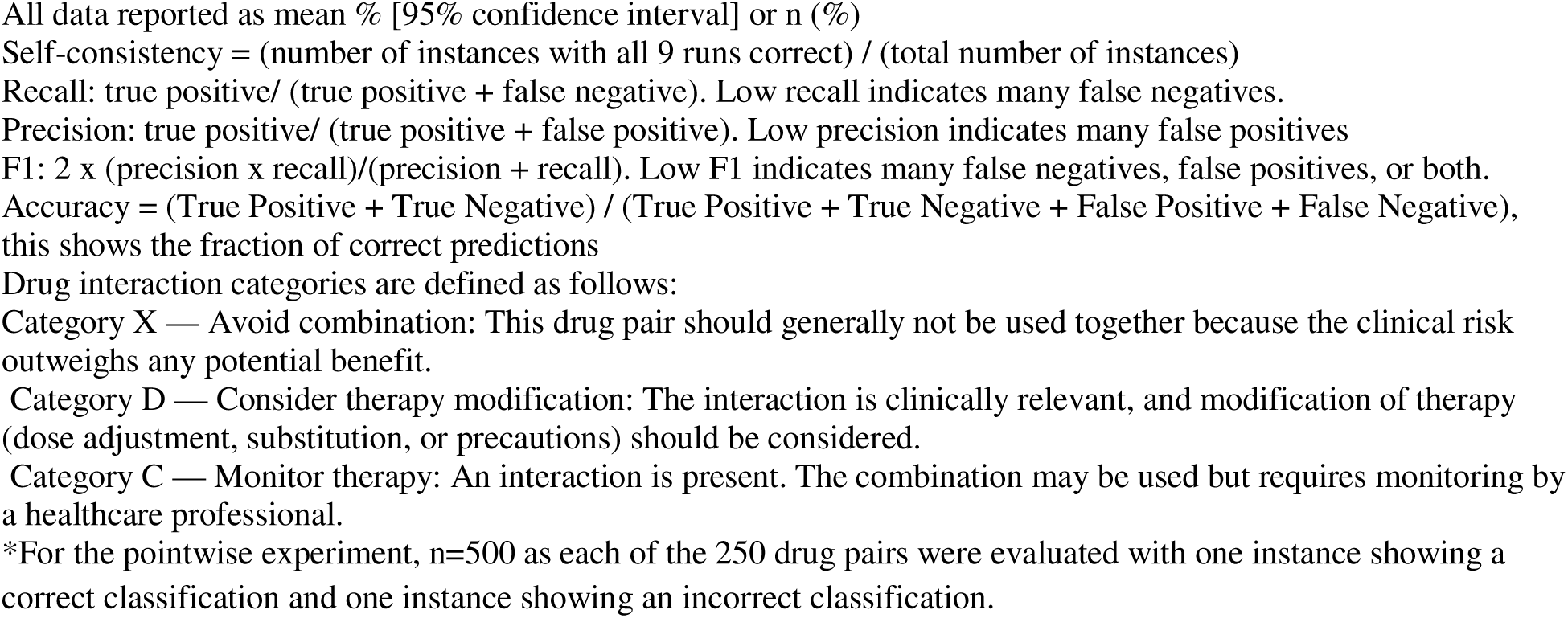
Results.

|  | **GPT-5-Chat**<br>**(n=250)** | **GPT-4o-mini**<br>**(n=250)** | **MedGemma-**<br>**27B**<br>**(n=250)** | **LLaMA3-70B**<br>**(n=250)** | **Qwen3-32B**<br>**(n=250)** | **DrugGPT**<br>**(n=250)** |
| --- | --- | --- | --- | --- | --- | --- |
| **Drug Pair Experiment (Pointwise)\*** |  |  |  |  |  |  |
| F1 | 80.2% [78.8, 81.7] | 54.5% [53.5, 55.7] | 42.2% [37.8, 46.7] | 59.7% [56.0, 63.4] | 69.4% [68.8, 70.1] | 45.0% [43.8, 46.2] |
| Precision | 81.3% [79.2, 83.5] | 65.7% [62.8, 68.8] | 51.8% [48.9, 54.4] | 60.8% [57.8, 63.9] | 59.6% [59.6, 59.6] | 50.1% [49.2, 51.0] |
| Recall | 80.7% [80.3, 81.1] | 46.7% [46.0, 47.5] | 38.2% [29.8, 46.6] | 61.7% [51.7, 71.7] | 64.1% [63.9, 64.4] | 40.9% [39.5, 42.4] |
| Accuracy | 80.6% [80.3, 80.8] | 61.0% [59.4, 62.6] | 50.1% [48.1, 52.0] | 59.8% [59.2, 60.5] | 66.7% [66.3, 67.1] | 50.1% [49.4, 50.8] |
| Self-Consistency Score | 371/500 (74.2) | 208/500 (41.6) | 155/500 (31.0) | 215/500 (43.0) | 307/500 (61.4) | 165/500 (33.0) |
| **3 Drug Combination Experiment (Pairwise)** |  |  |  |  |  |  |
| Accuracy | 93.6% [89.6, 95.6] | 86.6% [82.8, 91.2] | 84.9% [78.4, 87.6] | 81.5% [78.2, 84.6] | 84.0% [80.2, 87.4] | 93.7% [92.8, 94.5] |
| Self-Consistency Score | 234/250 (93.6) | 217/250 (86.6) | 212/250 (84.9) | 204/250 (81.6) | 210/250 (84.0) | 204/250 (81.6) |
| Category C (n=82) |  |  |  |  |  |  |
| Accuracy | 83.9% [73.2, 89.0] | 73.3% [63.4, 81.7] | 76.7% [59.8, 79.3] | 76.8 % [70.6, 82.5] | 75.5 % [68.9, 81.4] | 85.4% [83.5, 87.0] |
| Self-Consistency Score | 69/82 (84.1) | 60/82 (73.2) | 63/82 (76.8) | 63/82 (76.8) | 62/82 (75.6) | 49/82 (59.8) |
| Category D (n=78) |  |  |  |  |  |  |
| Accuracy | 99.2% [96.2, 100.0] | 92.3% [85.9, 97.4] | 89.2% [82.1, 96.2] | 89.0% [83.7, 93.9%] | 89.3% [78.2, 93.6] | 96.9% [95.2, 98.1] |
| Self-Consistency Score | 77/78 (98.7) | 72/78 (92.3) | 70/78 (89.7) | 69/78 (88.5) | 70/78 (89.7) | 70/78 (89.7) |
| Category X (n=90) |  |  |  |  |  |  |
| Accuracy | 97.8% [94.4, 100.0] | 93.8% [91.1, 98.9] | 88.6% [82.2, 95.6] | 79.3% [73.9, 84.1] | 87.2% [81.1, 94.4] | 98.6% [97.8, 99.4] |
| Self-Consistency Score | 88 (97.8) | 84 (93.3) | 80 (88.9) | 71 (78.9) | 78 (86.7) | 85 (94.4) |
| **4-6 Drug Combination Experiment (Listwise)** |  |  |  |  |  |  |
| Accuracy | 88.4% [84.4, 92.4] | 71.3% [66.4, 77.6] | 80.0% [75.2, 85.2] | 68.6% [63.2, 74.4] | 76.1% [70.8, 81.2] | 92.6% [91.6, 93.5] |
| Self-Consistency Score | 221/250 (88.4) | 178/250 (71.2) | 200/250 (80.0) | 172/250 (68.8) | 190/250 (76.0) | 205/250 (82.0) |
| Category C (n=81) |  |  |  |  |  |  |
| Accuracy | 78.1% [67.9, 86.4] | 65.0% [56.8, 76.5] | 73.8% [65.4, 84.0] | 58.3% [48.1, 67.9] | 65.6% [55.6, 75.3] | 86.7% [84.4, 88.8] |
| Self-Consistency Score | 63/81 (77.8) | 53/81 (65.4) | 60/81 (74.1) | 47/81 (58.0) | 53/81 (65.4) | 58/81 (71.6) |
| Category D (n=92) |  |  |  |  |  |  |
| Accuracy | 95.2% [91.3, 98.9] | 72.0% [62.0, 80.5] | 85.0% [77.2, 91.3] | 73.0% [63.0, 81.6] | 80.7% [71.7, 88.0] | 96.7% [95.9, 97.5] |
| Self-Consistency Score | 88/92 (95.7) | 66/92 (71.7) | 78/92 (84.8) | 67/92 (72.8) | 74/92 (80.4) | 79/92 (85.9) |
| Category X (n=77) |  |  |  |  |  |  |
| Accuracy | 91.3% [84.4, 96.1] | 77.2% [67.5, 87.0] | 80.5% [72.7, 89.6] | 74.3% [63.6, 84.4] | 81.8% [72.7, 90.9] | 93.9% [92.9, 94.8] |
| Self-Consistency Score | 70/77 (90.9) | 59/77 (76.6) | 62/77 (80.5) | 57/77 (74.0) | 63/77 (81.8) | 68/77 (88.3) |

All data reported as mean % [95% confidence interval] or n (%)
Self-consistency = (number of instances with all 9 runs correct) / (total number of instances)
Recall: true positive/ (true positive + false negative). Low recall indicates many false negatives.
Precision: true positive/ (true positive + false positive). Low precision indicates many false positives
F1: $2 \times (\text{precision} \times \text{recall}) / (\text{precision} + \text{recall})$ . Low F1 indicates many false negatives, false positives, or both.
Accuracy = $(\text{True Positive} + \text{True Negative}) / (\text{True Positive} + \text{True Negative} + \text{False Positive} + \text{False Negative})$ , this shows the fraction of correct predictions
Drug interaction categories are defined as follows:
Category X — Avoid combination: This drug pair should generally not be used together because the clinical risk outweighs any potential benefit.
Category D — Consider therapy modification: The interaction is clinically relevant, and modification of therapy (dose adjustment, substitution, or precautions) should be considered.

### Drug Pair Experiment (Pointwise)

Performance varied widely across models when classifying interaction categories for isolated drug pairs (**Table 3**). GPT-5-Chat demonstrated the strongest recall (80.7, 95% CI 80.3–81.1) and F1 score (80.2, 95% CI 78.8–81.7), indicating a greater ability to correctly identify interacting pairs. Other models had significantly worse performance with F1 scores of <70% for all other models tested. MedGemma-27B had the poorest performance, particularly in recall (38.2, 95% CI 29.8–46.6). Self-consistency was low for all models, with values ranging from 31.0-74.2%, consistent with earlier observations that LLM outputs often shift across repeated queries even when the underlying case remains unchanged. These findings echo prior studies showing instability in LLM clinical judgments, especially when tasks require precise multi-class classification.

### Three Drug Combination Experiment (Pairwise)

Accuracy improved considerably when models were asked to choose between two candidate drugs. DrugGPT achieved the highest overall accuracy (93.7, 95% CI 92.8–94.5), closely followed by GPT-5-Chat (93.6, 95% CI 89.6–95.6), and all models achieved an accuracy of >80%. Performance patterns were consistent across interaction categories, with models performing best on Category D and Category X interactions (accuracy 79.3-99.2%) compared to category C interactions (73.3-85.4%). Self-consistency values were higher than in the pointwise task (81.6-93.6%). This finding aligns with prior LLM evaluation frameworks where reduced answer space improves stability.

### Four-to-Six Drug Combination Experiment (Listwise)

Performance decreased in the more complex listwise setting, in which models were required to identify the interacting pair from a broader medication list. Accuracy ranged from 68.6% (LLaMA3-70B) to 92.6% (DrugGPT). All models showed their lowest accuracy in Category C interactions (58.3–86.7% across models), reflecting the difficulty of detecting moderate-risk interactions when combined with additional distractor drugs. Category X interactions generally showed higher accuracy across models (74.3–93.9%), although still lower than observed in the pairwise task.

As shown in **Figure 1**, the three models behave quite differently across the three DDI tasks. GPT-5-Chat shows the most even performance, staying relatively stable across the pointwise, pairwise, and listwise settings. DrugGPT performs best in the pairwise task and listwise task, where the model must identify an interacting pair from a larger set of drugs. However, while DrugGPT had higher accuracy in pairwise and listwise experiments, self-consistency was lower than other models, and the difference in accuracy was minimal compared to GPT-5-Chat, whereas the difference in accuracy in the pointwise experiment was much larger between GPT-5-Chat and all other models. **Figure 2** displays model self-consistency, with GPT-5-Chat having the highest self-consistency across all tasks.

**Figure 1.**
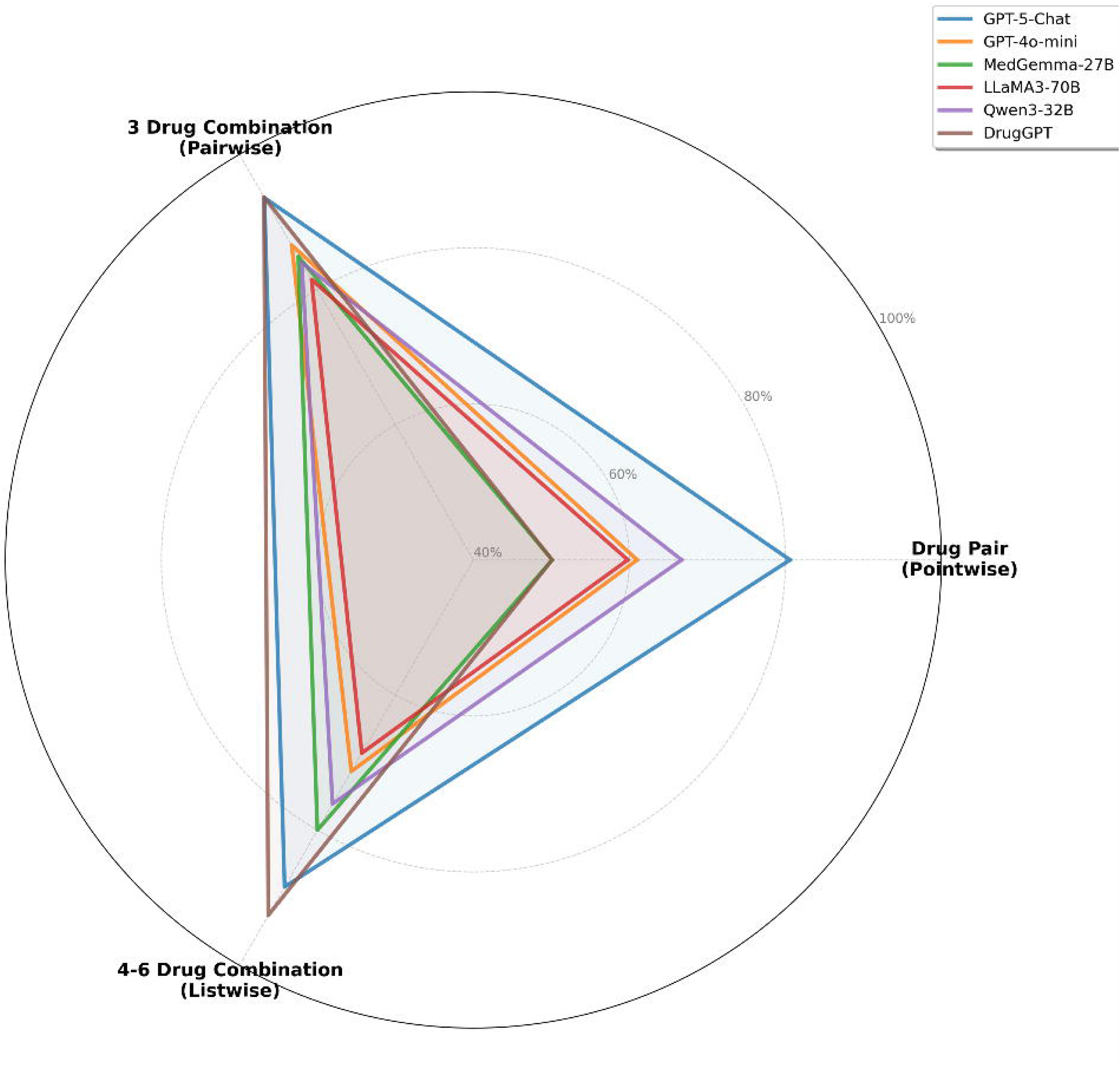
Radar Plot of Large Language Model Performance on Each Task Across all six models, the pairwise task is consistently easier than the pointwise task. This echoes patterns reported in other LLM evaluation work, where models tend to perform better when they can compare options directly rather than classify a single item in isolation. The results here follow the same trend: giving models two choices seems to reduce uncertainty and improve decision quality. Overall, the figure highlights that each model has a different “strength profile.”

**Figure 2.**
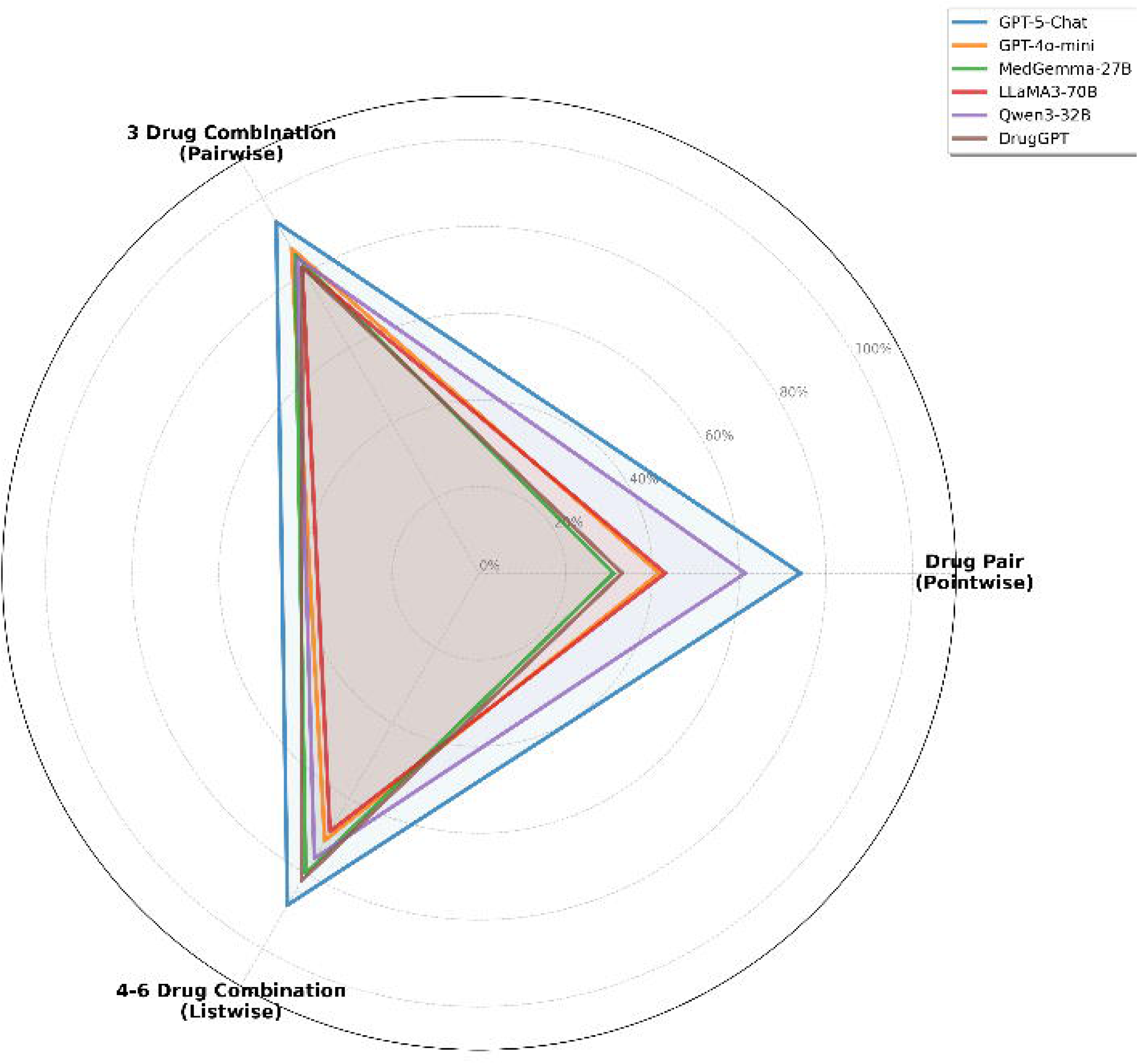
Radar Plot of Large Language Model Self-Consistency on Each Task Across all six models, the pointwise task resulted in lower self-consistency for both pairwise and listwise tasks.

## Discussion

This analysis marks the first time that a DDI dataset has been developed with robust validation of all DDIs by clinicians in addition to validation by a reputable drug information source (LexiDrug).^25^ Previously published DDI datasets were developed purely through extraction of DDIs from medical texts (e.g., DDI-Corpus) and lack the high-quality nature of a clinician-annotated dataset.^18,19,29–34^ Clinician annotated DDI datasets are rare and previously have only included annotations from experts with an informatics or biology background but have not included clinical annotations by in clinical medication use.^35^ A previous analysis did include an annotated dataset from a clinical pharmacist who selected relevant DDIs; however, this dataset only included DDIs for 5 medications (all macrolides or sodium-glucose transport 2 [SGLT2] inhibitors).^2^ The present study represents the first time that DDIs have been tested using a stepwise increase in task complexity. Previous reports have included drug pairs with a simple “yes/no” response to the question, “Do these drugs have an interaction?”^2,7,8,20^ These studies did not include additional clinical context and reported highly variable performance. Our study specifically included severity of DDIs for the drug-pair experiment and added increased complexity by providing a list of possible drugs to mimic information necessary to making clinical decisions in the a real-world setting. Several previous studies have included lists of medications (multiple medications per patient case) but did not consistently report the number of medications or compare LLM efficacy in identifying DDIs when the number of medications increased.^4,12,14,16,26^

Self-consistency scores declined across all models, which highlights that increasing task complexity leads to greater variability in model reasoning and lower reliability. This pattern mirrors observations in prior research, where reasoning stability deteriorates as the number of candidate options increases.^36,37^ Across the three experiments, several consistent patterns emerged. First, models performed better in structured decision settings with limited answer space, as seen in the pairwise task. Second, performance was consistently lower for moderate-risk Category C interactions, reflecting the subtlety and context dependence of these cases. Third, self-consistency remained imperfect even in tasks with high accuracy, indicating that correct answers do not guarantee stable reasoning. This finding aligns with prior work demonstrating that LLMs often produce confident but shifting outputs and that performance metrics alone can mask underlying instability.^38^

Several studies have evaluated LLM performance with real-world patient cases or medication profiles: one study demonstrated a low true positive rate and noted low agreement between GPT-3.5 and practicing pharmacists,^12^ while other studies have shown impressive DDI identification (ranging from 75.6-100%), but poor identification of the severity of the interaction.^14–16,20^ Few studies have evaluated the proficiency of LLMs in making clinical recommendations: one study found that ChatGPT, in response to sample textbook cases, achieved only 70% agreement with clinical pharmacists,^13^ and in two studies of real patient cases, ChatGPT provided appropriate recommendations for management of DDIs 88% and 61.5% of the time.^4,26^ Overall, LLMs tend to demonstrate acceptable performance in knowledge-based tasks, but significantly worse performance with complex or unstructured clinical tasks, which aligns with our results of decreasing performance as complexity increased.

This study has several limitations. The primary purpose of this study was to evaluate baseline performance or internal knowledge of LLMs for identifying DDIs, an important benchmarking task for the safe application of LLMs for medication management. However, it is known that prompt engineering and other LLM methods (e.g., knowledge graphs, multi-agent teams) can improve performance. However, even if such technology it exists, it is important to ensure safe performance on essential tasks, which this work can support. Second, while these datasets have an advantage in being clinician-developed to aid in clinical relevance and quality, they are relatively small by LLM standards. Finally, DDIs were clinically validated and correlated with drug interaction severity using a standard drug reference; however, additional references for DDIs may have slightly different categorizations for severity and existence of DDIs as new literature is available.

Ultimately, DDI identification was poor amongst all models, indicating worse performance than rules-based software upon which this database was built. Performance decreased as complexity increased, with identification of a DDI in cases of 4-6 medications only 69-93% of the time. This included not identifying category X DDIs (or severe interactions where the recommendation is to avoid the combination) in 6-26% of cases, where potential for patient harm is significant. Considering the average number of medications for an outpatient and inpatient regimen is 4-7 and 7-15, respectively, and the known positive association of adverse drug events with an increasing number of medications, there is concern for using LLMs to identify DDIs, due to the worsened performance with increasing complexity of medication regimen.^39–42^ Unidentified DDIs included interactions such as enoxaparin and alteplase (increased bleeding risk), sulfamethoxazole-trimethoprim and spironolactone (hyperkalemia), dronedarone and azithromycin (QTc prolongation), and phenytoin and apixaban (decreased effectiveness of apixaban). These few examples of undetected DDI pose high risk situations for potentially harmful adverse drug events and would have likely resulted in medication modification or increased monitoring with the initiation of therapy. Poor precision risks identification of a DDI that does not exist. In clinical practice, this could lead to alterations in treatment regimens resulting in worsened clinical outcomes and unnecessary monitoring. Again, it is important to realize that rules-based computer software has 100% performance in this domain. If future LLMs could identify DDIs at this performance level and provide more individualized recommendations (e.g., two sedative agents may be used together for alcohol withdrawal and do not necessitate a DDI warning for sedation), this would be a tangible improvement to present day software. Until then, safety risks potentially outweigh any benefits.

## Conclusion

The LLMs had moderate performance on three different DDI tasks, with performance generally decreasing with increased complexity (both increasing number of medications and requiring the LLM to specify the interaction severity). Further model improvement is necessary for routine use for identification of clinically relevant DDIs.

## Data Availability

All data produced in the present study are available upon reasonable request to the authors

https://github.com/sikora07/AIChemist

## Notes

**Conflicts of Interest:** The authors have no conflicts of interest.

### Competing Interest Statement

The authors have declared no competing interest.

### Funding Statement

Funding through Agency of Healthcare Research and Quality for Dr. Sikora was provided through R21HS028485 and R01HS029009.

### Summary of Updates

Updated to include a sixth LLM, DrugGPT, and 3 additional authors.

