## Supplementary figures and images for "Drug-drug interaction identification using large language models"

### Supplementary Appendix

Supplementary Appendix

Table of Contents

[TRIPOD+LLM Checklists	2](#_Toc228516326)

#### TRIPOD+LLM Checklists

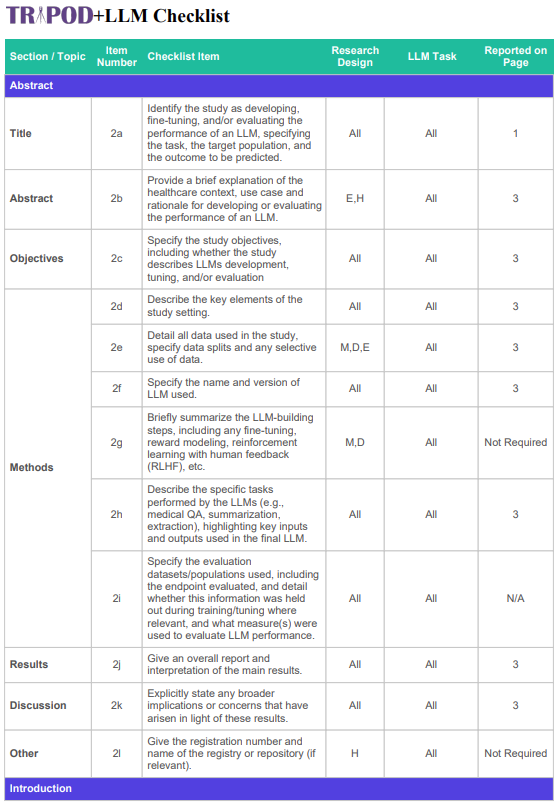

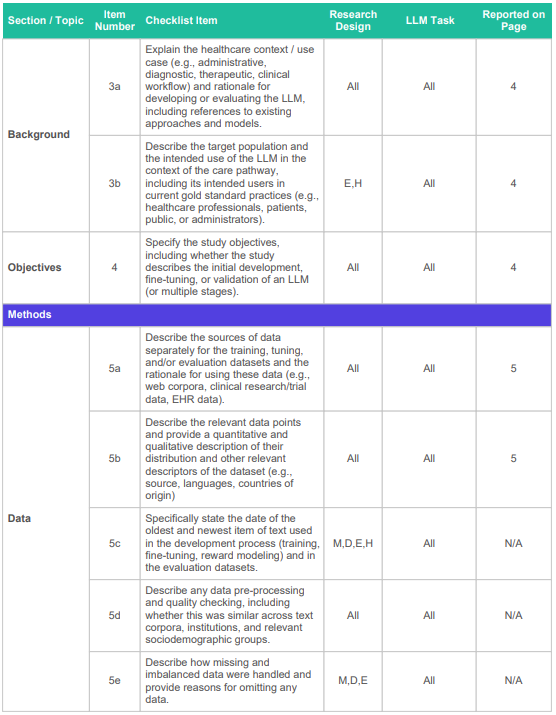

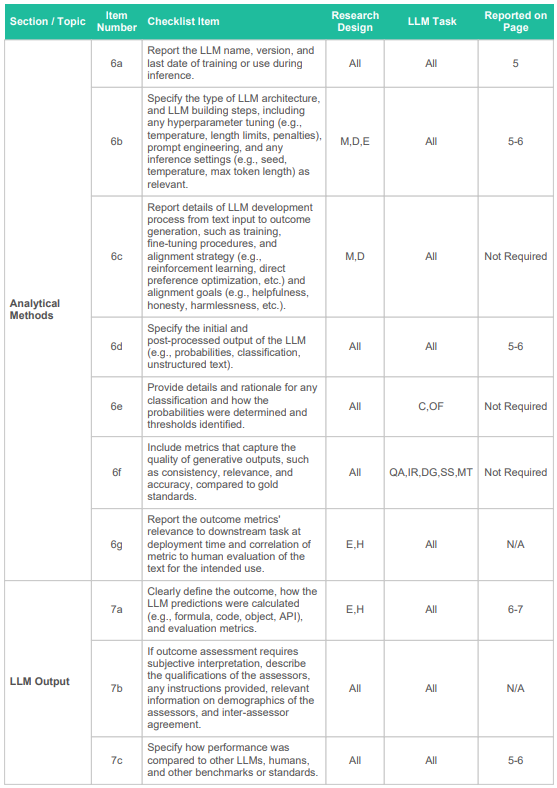

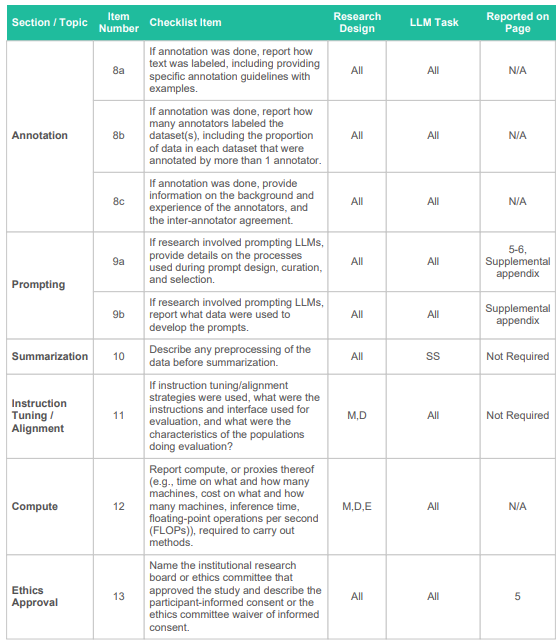

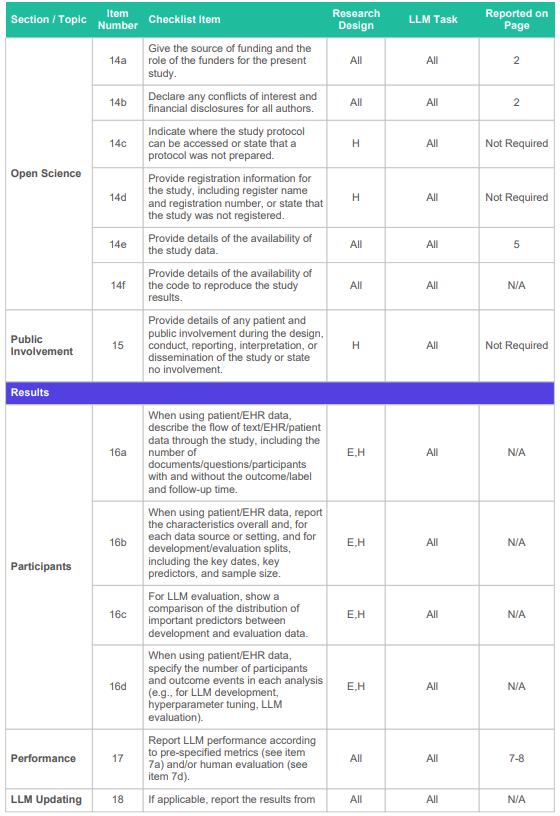

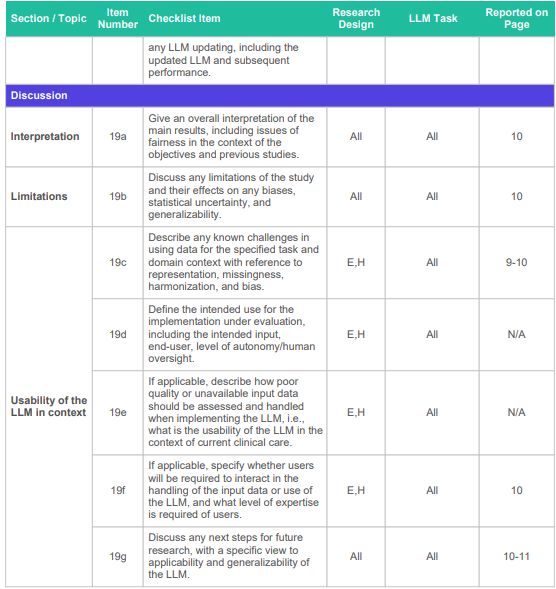
